# Epidemiological and clinical characteristics of COVID-19 in adolescents and young adults

**DOI:** 10.1101/2020.03.10.20032136

**Authors:** Jiaqiang Liao, Shibing Fan, Jing Chen, Jianglin Wu, Shunqing Xu, Yuming Guo, Chunhui Li, Xianxiang Zhang, Chuansha Wu, Huaming Mou, Chenxi Song, Feng Li, Guicheng Wu, Jingjing Zhang, Lian Guo, Huawen Liu, Jinglong Lv, Lixin Xu, Chunhui Lang

## Abstract

**Background:** Adolescents and young adults might play a key role in the worldwide spread of Coronavirus Disease 2019 (COVID-19), because they are more involved in overseas studying, business, working, and travelling. However, the epidemiological and clinical characteristics of them are still unknown.

**Methods:** We collected data of 46 confirmed COVID-19 patients aged 10 to 35 years from the study hospital. The demographics, epidemiological, and clinical data were collected. Several key epidemiological parameters, the asymptomatic cases and transmission to their family members and the clinical characteristics at admission, and during treatment were summarized.

**RESULTS:** Of 46 confirmed patients, 14 patients (47.3%) were aged from 10 to 24 years, and 24 (52.7%) patients were male. The mean incubation period for symptomatic cases was 6.6 days (95% confidence interval (CI) 4.4 - 9.6). The median serial interval was 1.9 days (95% CI 0.4 - 6.2). Three of asymptomatic cases showed the transmission to their family members. Only 1 patient was identified as severe cases at admission. The common symptoms at admission were dry cough (34, 91.0%), and fever (29, 69.0%). Nearly 60% of the patients had showed ground-glass opacity by chest CT findings. Three patients developed acute kidney injury during treatment. Majority of patients (78.3%) were discharged by the end of the follow-up.

**Conclusions:** The adolescent and young adult patients of COVID-19 had a long incubation period, and a short serial interval. The transmission to their family contactors occurred in asymptomatic cases. Few of the study patients have developed complications during treatment.

## Introduction

The coronavirus disease (COVID-19), a newly emerging infectious pneumonia with unknown causes, was originated in Wuhan, Hubei Province, China since December, 2019. Since its special onset time (during the Chinese spring festival), the incidences of COVID-19 were rapidly reported across China^1^. Early epidemiological and clinical studies depicted that the majority of the patients were middle-aged, or elder individuals, with a mean incubation period of 5.2 days (range 0-14 days), and a serial interval of 7.5 days (95% CI 2-17 days)^2-4^. The most common symptoms were fever, cough, and fatigue^3-5^. Most of the patients presented the abnormalities of chest CT-findings such as ground-glass opacity, and bilateral patchy shadowing^4-6^. The patients aged above 65 years were more likely to be severe cases and developed severe acute complications such as pneumonia, Acute Respiratory Distress Syndrome (ARDS), shock, and acute cardiac injury during treatment^3-5^. These studies provided essential evidence to guide the early medical screen, diagnosis of COVID-19 cases, isolating of the suspected individuals, and clinical treatments. However, with the rapid progress of COVID-19, many new characteristics have emerged, which need update more evidence. The most important point is that there has been increasing younger patients were confirmed across China. One study from Chinese Center for Disease Control and Prevention indicated that 4168 (9%) of the patients through February 11, 2020 were aged younger than 30 years ^7^. In addition, outbreaks of COVID-19 were reported worldwide. It is suspected that their first-generation cases were imported from China^8-10^. Several countries, such as South Korea, Japan, and Italy are experiencing a sharp increase in incident COVID-19 confirmed cases. Younger individuals were more likely to be carriers of COVID-19 across counties, since they were more likely involved in overseas study, business, work and travel. For example, in South Korea, 178 out of 431 confirmed cases, which were publicly announced in the website of Ministry of Health and Welfare, were aged ≤35 years through March 1, 2020^11^. However, as far as we know, no study has been specifically conducted to research the epidemiological and clinical characteristics in younger patients of COVID-19. It is an essential step to prevent the worldwide epidemic of COVID-19 in the future.

In this study, basing on a retrospective case series data, we aimed to estimate the key epidemiological characteristics and describe the clinical symptoms, treatments, and hospital outcomes for COVID-19 patients in adolescents and young adults.

## Methods

### Study Design and Participants

In this study, we defined the adolescent as 10-24 years of age and young adult as 25-35 years of age according to the World Health Organization’s definition. We retrospectively reviewed the medical records of confirmed COVID-19 cases aged from 10 to 35 years who were hospitalized in Chongqing Three Gorges Central Hospital of Chongqing University from January 25, 2020 to February 18, 2020.

### Data Collection

Epidemiological data were collected using standardized questionnaire through face-to-face or telephone interviewing with patients or their family members. We firstly collected the demographic and social-economic information such as height and weight, educational level, and behavioral characteristics such as smoking, alcohol consumption, and physical activities. Then, we investigated the exposure date and types for each patient during the 1 month before the date of symptoms onset. For those who resided in Wuhan, we further collected the histories of exposing to the Huanan Seafood wholesale market or other similar market. We collected the earliest date of symptoms onset and the specific symptoms. For those who were the first case of developing symptoms out of family (index patients), we further interviewed their family contactors with exposure histories with index patient, date of symptoms onset, date of medical visit, and date of confirmation. The clinical information for confirmed cases were abstracted from medical records. We collected several key information of date including date of clinical symptoms onset, date of primary visit to health facilities, and date of confirmation. The typical clinical symptoms and the data of chest CT scan for each case were collected at the admission and during the treatment. The medical histories and treatments such as antiviral therapy, antimicrobial therapy, corticosteroid therapy, and respiratory support were simultaneously recorded. We further collected the data of complications for cases during treatment. The ARDS was defined as the interim guidance of WHO for novel coronavirus, and acute kidney injury was defined on the basis of the highest serum creatinine level or urine output criteria according to the kidney disease improving global outcomes classification^12 13^. Cardiac injury was defined if the serum levels of cardiac biomarkers (eg, troponin I) were above the 99th percentile upper reference limit or new abnormalities were shown in electrocardiography and echocardiography. The clinical outcomes (discharge, still treatment, or death) were consistently observed until the date of February 23, 2020. The epidemiological data were inputted by Epidata with double checking. To ensure the accuracy of the clinical data, Two researchers also independently reviewed the electronic medical records.

### Laboratory confirmation and tests

The criteria of diagnosis for COVID-19 cases was based on national recommendation of the New Coronavirus Pneumonia Prevention and Control Program (6^th^ edition) ^14^. Briefly, the throat swab samples or lower respiratory tract were collected and processed at the department of clinical laboratory of study hospital. Then, the 2019-nCoVRNA were extracted from the patients who were suspected of having 2019-nCoVinfection. Finally, the throat swabs were placed into a collection tube with 150 μL of virus preservation solution, and total RNA was extracted within 2 hours using there spiratory sample RNA isolation kit (Suzhou Tianlong Biotechnology Co. Ltd,Roche’s COBASZ480). A Reverse Transcription-Polymerase Chain Reaction (RT-PCR) assay with a cycle threshold value (Ct-value) less than 37 was defined as a positive test result. Asymptomatic cases were defined as those who presented positive results by conducting the nucleic acid test of COVID-19, and had no elevated temperature measured or self-reported fever and no gastrointestinal or respiratory symptoms such as cough and sore reported by physicians at admission. To confirm the validity, we further conducted a face-to face or telephone interview to collect information of symptoms before 2 weeks of admission for each asymptomatic case. For each patient, the laboratory tests were performed at admission, which included routine blood tests, serum biochemistry, and coagulation function.

### Statistical analysis

We described the differences of demographic factors, symptoms at admission, comorbidities, and chest CT findings (10-24y, and 25-35y). We summarized the distribution of laboratory findings for cases using median and interquartile range among groups of total cases, 10-24y, and 25-35y. We defined the incubation period as the time interval from the date of exposure to the date of symptoms onset. We included two types of patients who could recalled the exact date of travelling to Wuhan or contacting with other confirmed cases. To ensure the accuracy, those who have reported more than one exposure sources or contacting periods over 3 days were excluded. We used a parametric survival analysis model with Weibull distribution to estimate the distribution of incubation period. Since the asymptomatic cases at first medical visit could develop symptoms during the follow up, we firstly treated the incubation period for asymptomatic cases as right-censored data (from the date of exposure to the date of first medical visit) and performed the estimations. Then, we excluded the asymptomatic cases and repeated the estimations. We defined the family clustered events as those who were the first cases of developing symptoms in their family (index patient), and their family members (secondary cases) had a clear contact history to the index patient and had no other potential infection source. We used the date of symptoms onset to measure the date of illness onset. We defined the serial interval as the time interval from the dates of illness onset between index case and the secondary cases. We used a parametric survival analysis model with gamma distribution to estimate the distribution of serial interval. We further calculated the time interval from the date of symptoms onset to the date of first medical visit using parametric survival model with Weibull distribution. We finally compared the differences of treatments, days of persisting fever during treatment, days of transforming to negative results by COVID-19 nucleic acid tests into negative during treatment, and prognosis outcomes by different age groups.

We performed the summaries and significance tests by SAS 9.4, The parametric survival analyses were conducted by R 3.1.1, The statistically significant level was defined as 0.05 with a 2-side test.

### Ethical approval

Data collection and analyses of cases were approved by the institutional ethnic board of three gorges hospital affiliated of Chongqing University (No.2020-7(论)). Since the epidemiological interview for the study cases is a part of a continuing public health outbreak investigation determined by the National Health Commission of the People’s Republic of China and the identified information was deleted. The individual consent was exempted.

## Results

We totally included 46 hospital diagnosed COVID-19 patients (Table 1). Majority of them were young adults (n=32), and the rest were adolescents (n=14). The main exposure types of patients included contacting with other confirmed cases (22,47.8%) or residing in Wuhan (19, 41.3%). Compared with young adult, adolescent was more likely to reside or travel to Wuhan. Half of the patient were men (24, 52.2%), had a normal BMI (24, 52.2%), and never conducted physical activity (23, 50.0%). Compared with young adult, adolescent was less likely to be overweight/obesity, smoking, never conducting physical activity, and never drinking alcohol. Few of patients had 1 or more medical disease histories (6, 13.0%). The specific medical diseases included obesity (1), diabetes (1), Chronic obstructive pulmonary disease (1), hyperthyroidism (1), kidney stones (1), and arthrolithiasis (1). Only one (2.2%) patient was severe case, and 4 (8.7%) were asymptomatic at admission. The most common symptoms at admisson were dry cough (34, 81.0%), fever (29, 69.1%), and expectoration (16, 38.1%). The less common symptoms included headache, fatigue, pharyngalgia, chest pain, anorexia, myalgia, dizziness, diarrhea, nausea, and shortness of breath. The common pathological changes of chest CT findings were ground-glass opacity (29, 63.0%), and bilateral patchy shadowing (12, 26.1%). Compared with young adults, no severe cases and higher odds of asymptomatic cases (14.3% vs 6.3%) were observed in adolescent patients. Fewer adolescent patients reported fever, headache, and fatigue at onset illness. Only 7 (50.0%) of adolescent patients showed ground-glass opacity for chest CT scanning, compared with 22 (68.8%) of that in young adult. We displayed the typical patterns of chest CT scan for adolescents and young adults in Figure 1.

**Table 1.**
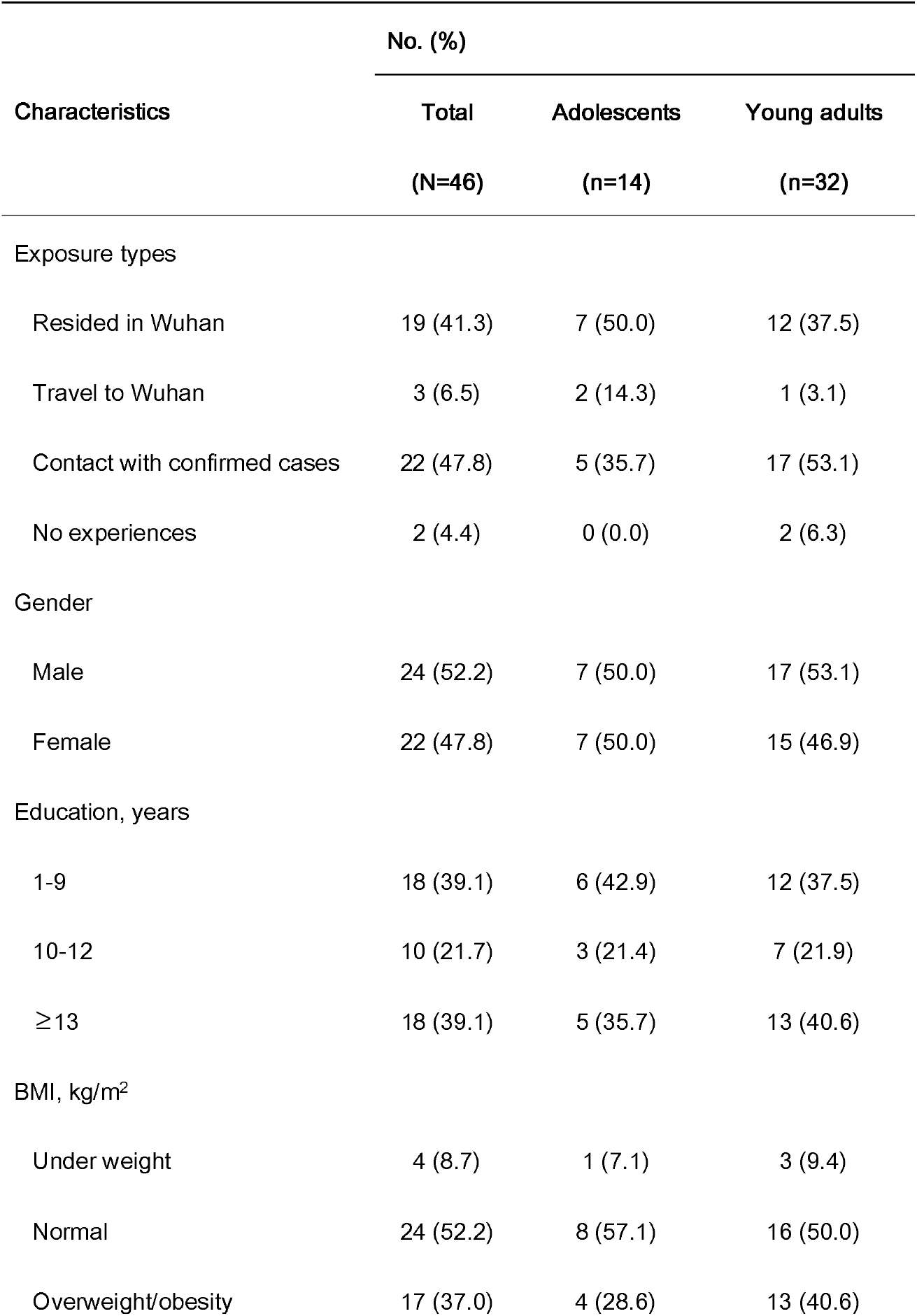
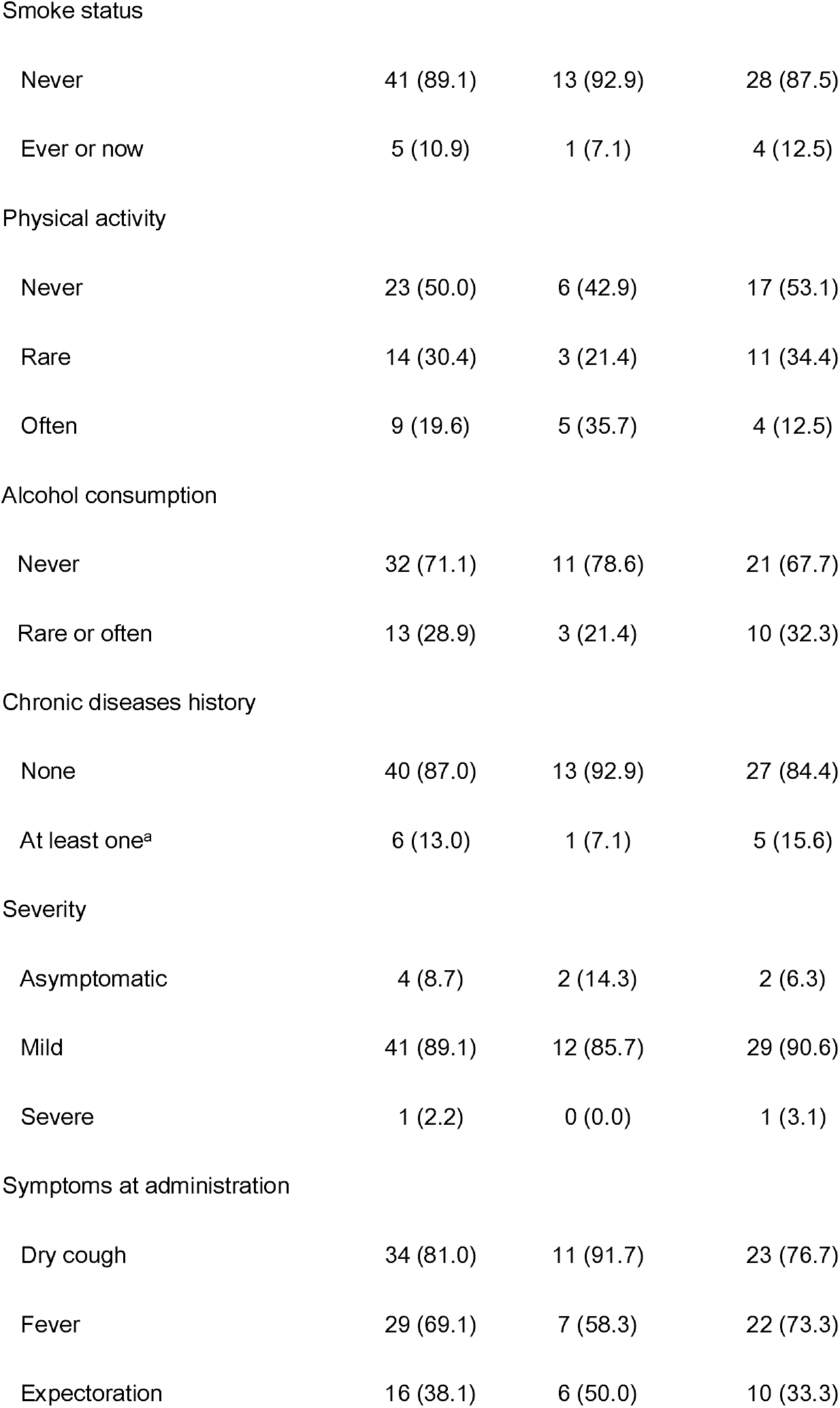
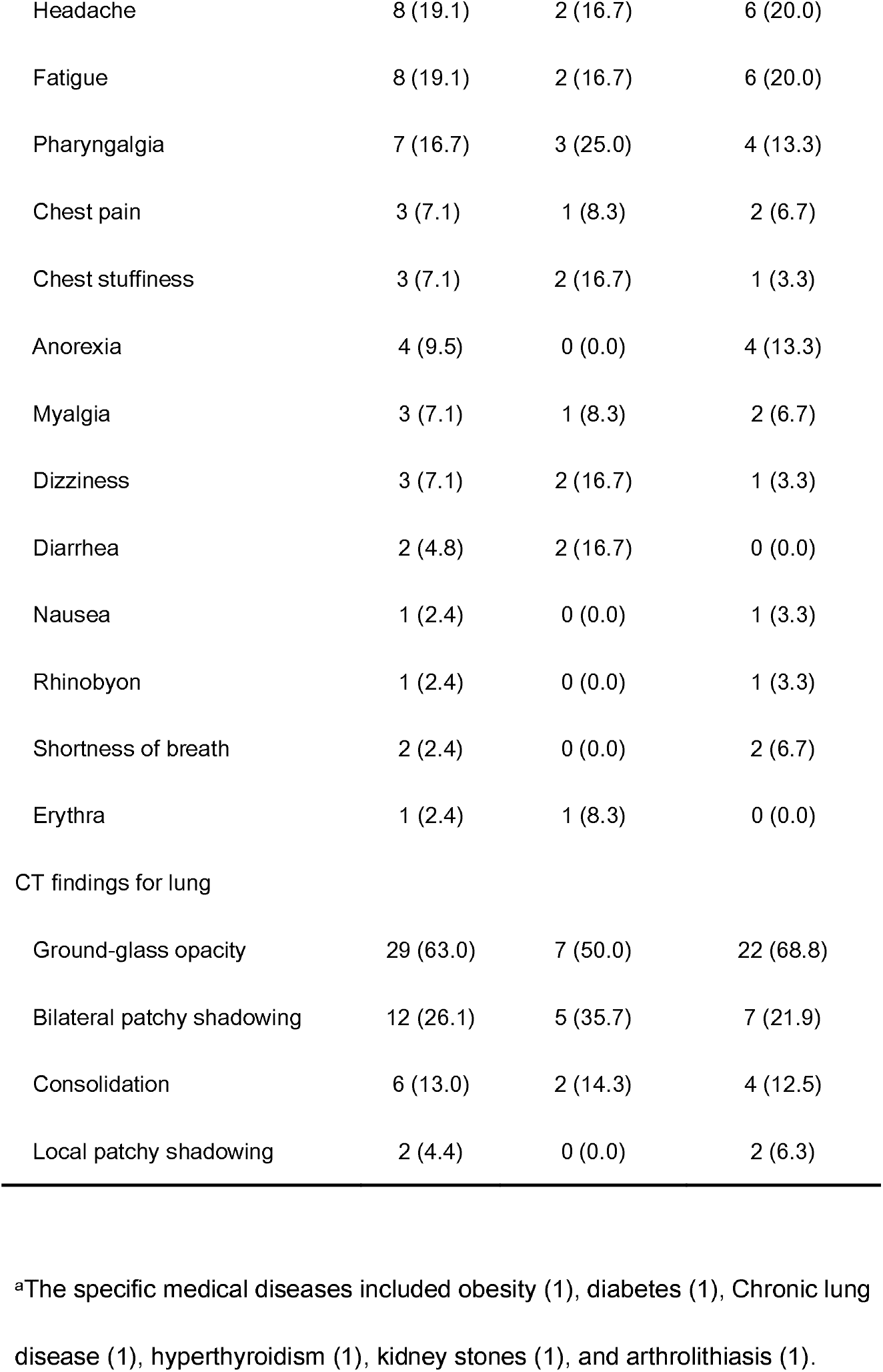
Baseline characteristics of study patients infected with COVID-19.

**Figure 1.**
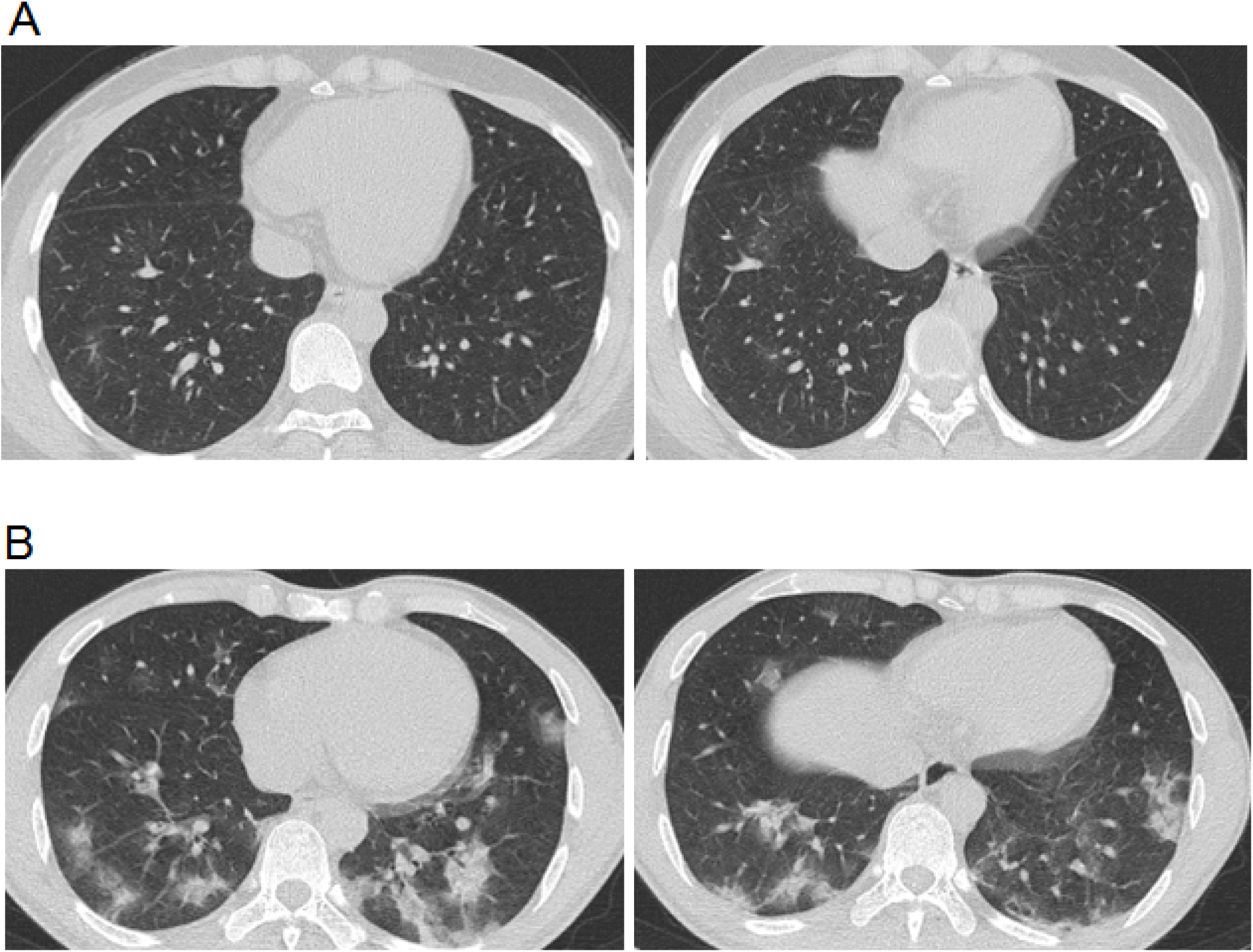
Chest computed tomographic (CT) images for study patients infected with COVID-19. Panel A depicted the chest CT images for a patient aged 21 years on day 10 after illness onset, and Panel B depicted the chest CT images for a patient aged 33 years on day 14 after illness onset.

We recorded family clustered events from 6 symptomatic cases at admission (Figure 2). Among 14 patients who provided exact date of travelling to Wuhan or contacting with other confirmed cases, the estimated median incubation period was 8.3 days (95% CI 5.0 −13.4) (Figure 3 Panel A). The estimated 95^th^ percentile of incubation period could reach as long as 24.8 days (95% CI 14.9 - 47.6). After excluding 3 asymptomatic cases, the estimated median incubation period decreased to 6.6 days (95% CI 4.4 - 9.6) (Figure 3 Panel B). The estimated 95^th^ percentile of incubation period decreased to 14.8 days (95% CI 10.4 - 22.0). According to the definition of serial interval, we only included 12 secondary cases who provided exact dates of illness onset from 6 families with symptomatic index cases to estimate the distribution of serial interval (Figure 3 Panel C). The estimated median serial interval was 1.9 days (95% CI 0.4 - 6.2). The estimated 95^th^ percentile of serial interval could reach as long as 28.6 days (95% CI 10.6 −76.9). Based on 42 symptomatic cases, we estimated the median days from symptom onset to first medical visit to be 1.4 days (95% CI 0.8 - 2.4) (Figure 3 Panel D). The estimated 95^th^ percentile days from symptom onset to first medical visit was 13.2 days (95% CI 8.3 - 20.9).

**Figure 2.**
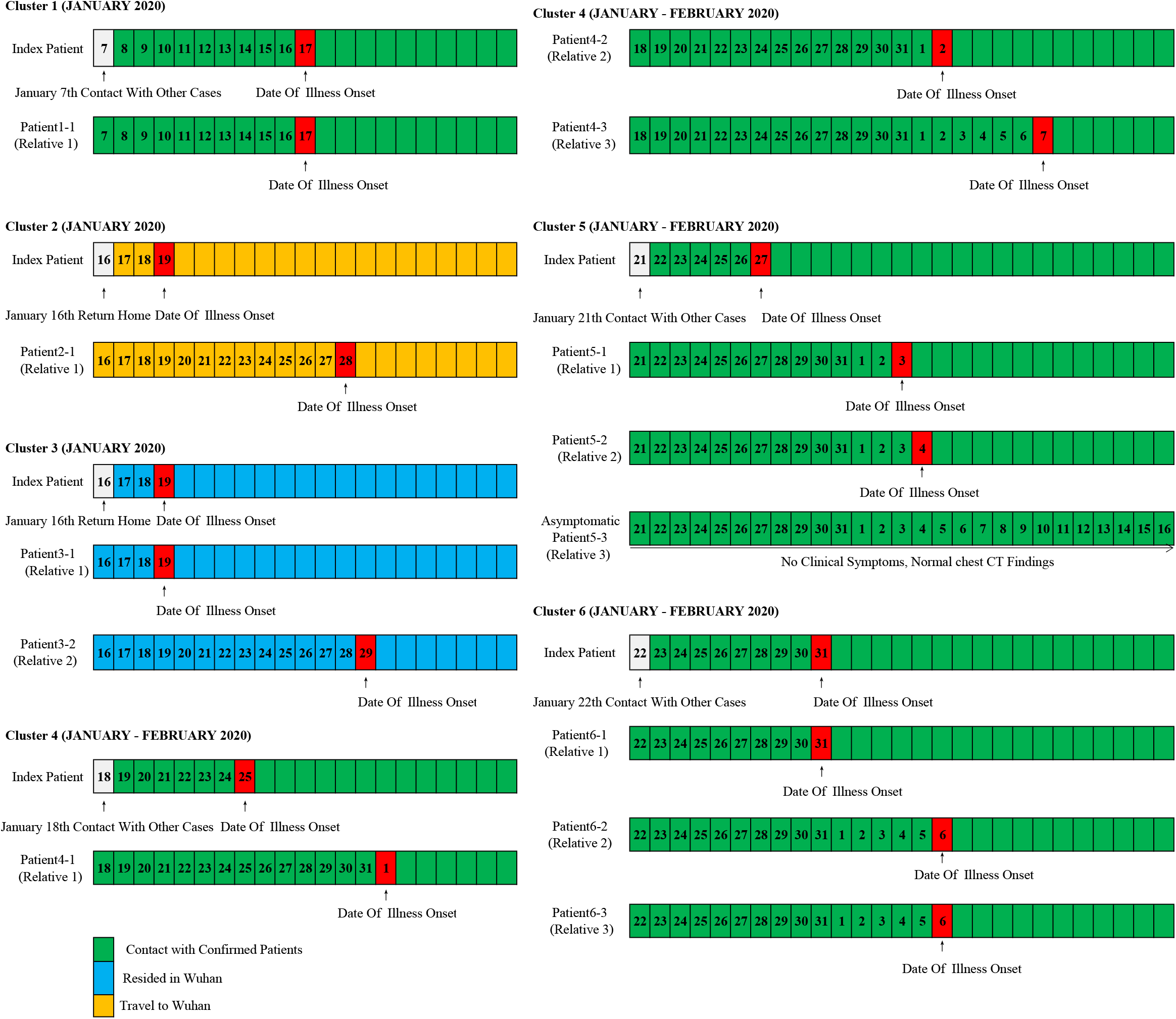
Information on exposures and dates of illness onset in 6 symptomatic cases and their family close contactors. Numbers in boxes are calendar dates. Data from the 12 secondary cases (close contactors were defined as those who had clear exposure to only one index case and had no other potential source of infection) were used to estimate the distribution of serial interval.

**Figure 3.**
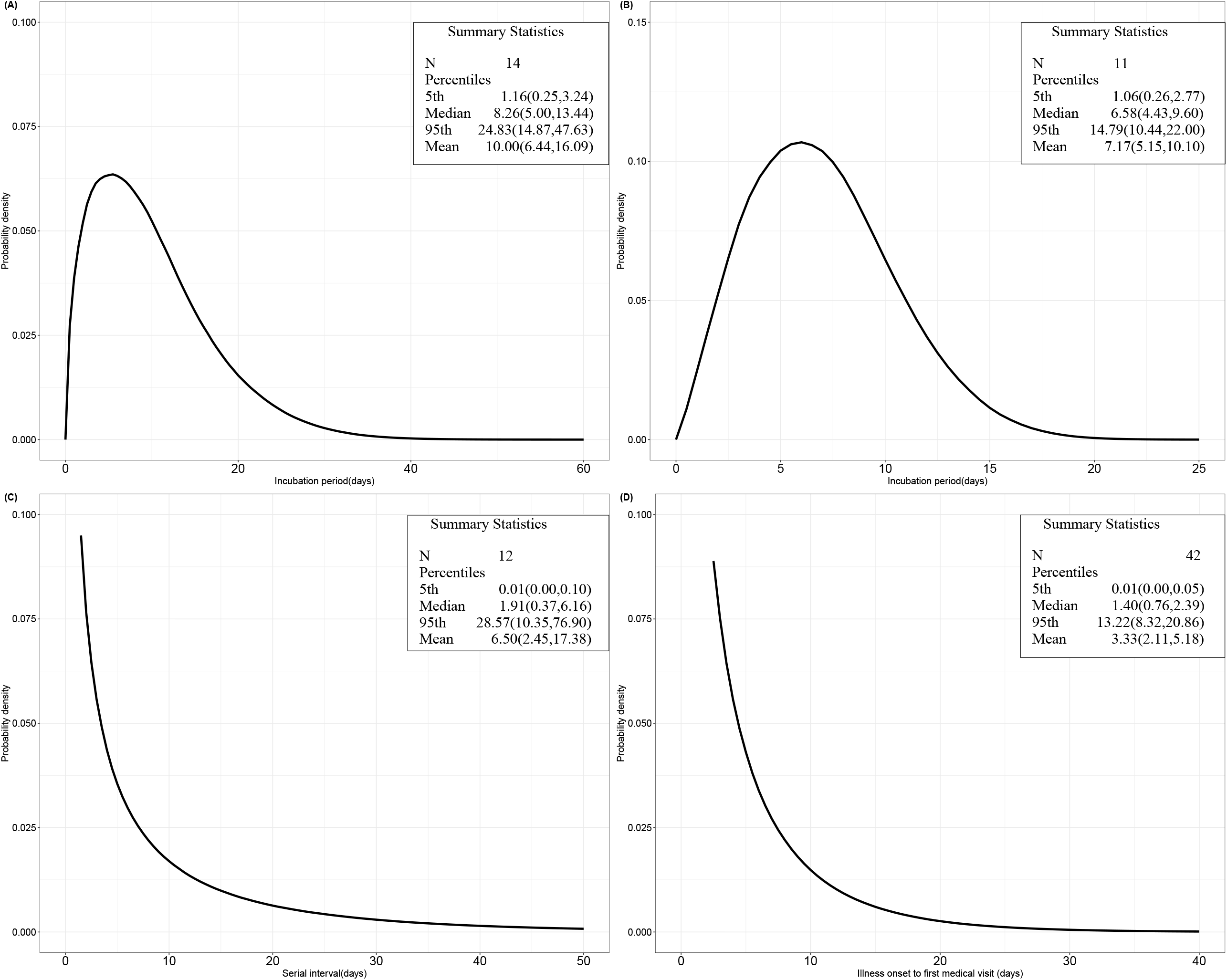
Key distributions of epidemiological characteristics for study patients. The estimated incubation period distribution for symptomatic cases and asymptomatic cases truncated at hospitalization is depicted in Panel A. The estimated incubation period distribution only for asymptomatic cases is depicted in Panel B. The estimated serial interval distribution is depicted in Panel C. The estimated distributions of times from illness onset to first medical visit is depicted in Panel D.

On admission, 10 (21.7%) patients were leucopenia (white blood cell count < 4 × 10^9^/L), and 29 (63.0%) patients were lymphopenia (Table 2). Ten patients (21.7%) had decreased levels of platelet count (< 150× 10^9^/L). The other elevated levels of laboratory indicators for study patients were lactate dehydrogenase (9, 19.6%), C-reactive protein (9, 19.6%), D-dimer levels (7, 15.2%), alanine aminotransferase (7, 15.2%), and total bilirubin (7, 15.2%). Numerous differences were observed in laboratory findings between adolescent and young adult patients. For example, 8 (25.0%) young adult patients presented elevated levels of c-reactive protein, while only 1 adolescent patient showed the similar pattern.

**Table 2.** Laboratory findings of study patients on admission. Values are medians (interquartile ranges) unless stated otherwise.

| Variables | Total<br>(N=46) | Adolescents<br>(n=14) | Young adults<br>(n=32) |
| --- | --- | --- | --- |
| White blood cell count ($\times 10^9/L$) | 5.0 (4.1- 6.7) | 5.4 (4.5- 6.7) | 4.8 (3.9- 6.7) |
| <4 (No(%)) | 10.0 (21.74) | 1.0 (7.1) | 9 (28.1) |
| >10 (No(%)) | 2.0 (4.4) | 0 (0.0) | 2 (6.3) |
| Neutrophil count ($\times 10^9/L$) | 3.4 (2.5- 4.4) | 3.8 (3.1- 5.6) | 3.2 (2.4- 4.4) |
| Lymphocyte count ($\times 10^9/L$) | 1.3 (1.0- 1.8) | 1.4 (1.1- 2.3) | 1.3 (0.9- 1.8) |
| <1.5 (No(%)) | 29 (63.0) | 8 (57.1) | 21 (65.6) |
| Platelet count ($\times 10^9/L$) | 192.5 (156.0- 237.0) | 183 (156.0- 227.0) | 194.5 (152.0- 248.0) |
| <150 (No(%)) | 10 (21.7) | 2 (14.3) | 8 (25.0) |
| Haemoglobin (g/L) | 139.5 (130.0- 151.0) | 147 (133.0- 151.0) | 137.5 (128.5- 150.5) |
| Prothrombin time (S) | 11 (10.6- 11.4) | 11.25 (10.7- 11.9) | 11 (10.5- 11.1) |
| Activated partial thromboplastin time (S) | 27 (25.9- 30.3) | 28.2(26.1- 30.4) | 26.9 (25.0- 30.3) |
| D-dimer (mg/L) | 0.3 (0.2- 0.4) | 0.2 (0.1- 0.3) | 0.3 (0.2- 0.4) |
| $\geq 0.5$ (No(%)) | 7 (15.2) | 3 (21.4) | 4 (12.5) |
| Alanine aminotransferase (U/L) | 17.9 (11.6- 32.5) | 16.3 (7.1- 24.3) | 20 (11.7- 32.7) |
| >40 (No(%)) | 7 (15.2) | 1 (7.2) | 6 (18.8) |
| Aspartate aminotransferase (U/L) | 18.3 (14.5- 26.9) | 16.6 (12.4- 23.6) | 19.3 (15.8- 28.6) |
| >40 (No(%)) | 3 (6.5) | 0 (0.0) | 3 (9.4) |
| Total bilirubin, $\mu\text{mol/L}$ | 8.7 (5.9- 14.6) | 9 (5.9- 24.8) | 8.3 (6.1- 13.5) |
| >17.1 (No(%)) | 7 (15.3) | 4 (28.6) | 3 (9.4) |
| Blood urea nitrogen, mmol/L | 3.4 (2.6- 4.8) | 3.9 (2.6- 4.9) | 3.15 (2.6- 4.6) |
| Creatine ($\mu\text{mol/L}$) | 61.5 (52.0- 79.0) | 62.5 (53.0- 81.0) | 61.5 (51.5- 77.0) |
| Creatine kinase (U/L): | 57 (41.0- 73.0) | 53 (42.0- 73.0) | 57.6 (36.0- 74.9) |
| $\geq 200$ (No(%)) | 2 (4.4) | 0 (0.0) | 2 (6.3) |
| Lactate dehydrogenase (U/L): | 195.5 (145.0- 240.0) | 180 (152.0- 220.0) | 200 (144.0- 244.0) |
| $\geq 250$ , (No(%)) | 9 (19.6) | 2 (14.3) | 7 (21.9) |
| Procalcitonin, ng/ml | 0.03 (0.0- 0.1) | 0.04 (0.0- 0.1) | 0.03 (0.0- 0.1) |
| $\geq 0.1$(No(%)) | 2 (4.4) | 0 (0.0) | 2 (6.3) |
| C-reactive protein, mg/L | 2.6 (0.8- 9.4) | 3.2 (0.3- 5.6) | 3.0 (1.0- 10.0) |
| $\geq 10$ (No(%)) | 9 (19.6) | 1 (7.1) | 8 (25.0) |

The treatments and prognosis outcomes were summarized in Table 3. During the treatment periods, all patients received antiviral therapy, 39 (84.8%) patients received oxygen inhalation, and 43 (93.5%) patients received interferon alpha inhalation. Few patients (5, 10.9%) received antifungal treatment. Three (6.5%) patients have developed acute kidney injury during the treatment. The estimated median days from the date of admission to the date of consecutively negative results for COVID-19 nucleic acid tests were 12.6 days (95% CI 11.2 - 14.1). The median days of persistent fever during admission were 5 days (IQR 1-8). Until the date of 25^th^ February 2020, 36 patients had been discharged, 10 patients were hospitalized, and no patients were died. Compared with young adult, adolescent patients received less therapy of oxygen inhalation and had shorter days of persistent fever.

**Table 3.**
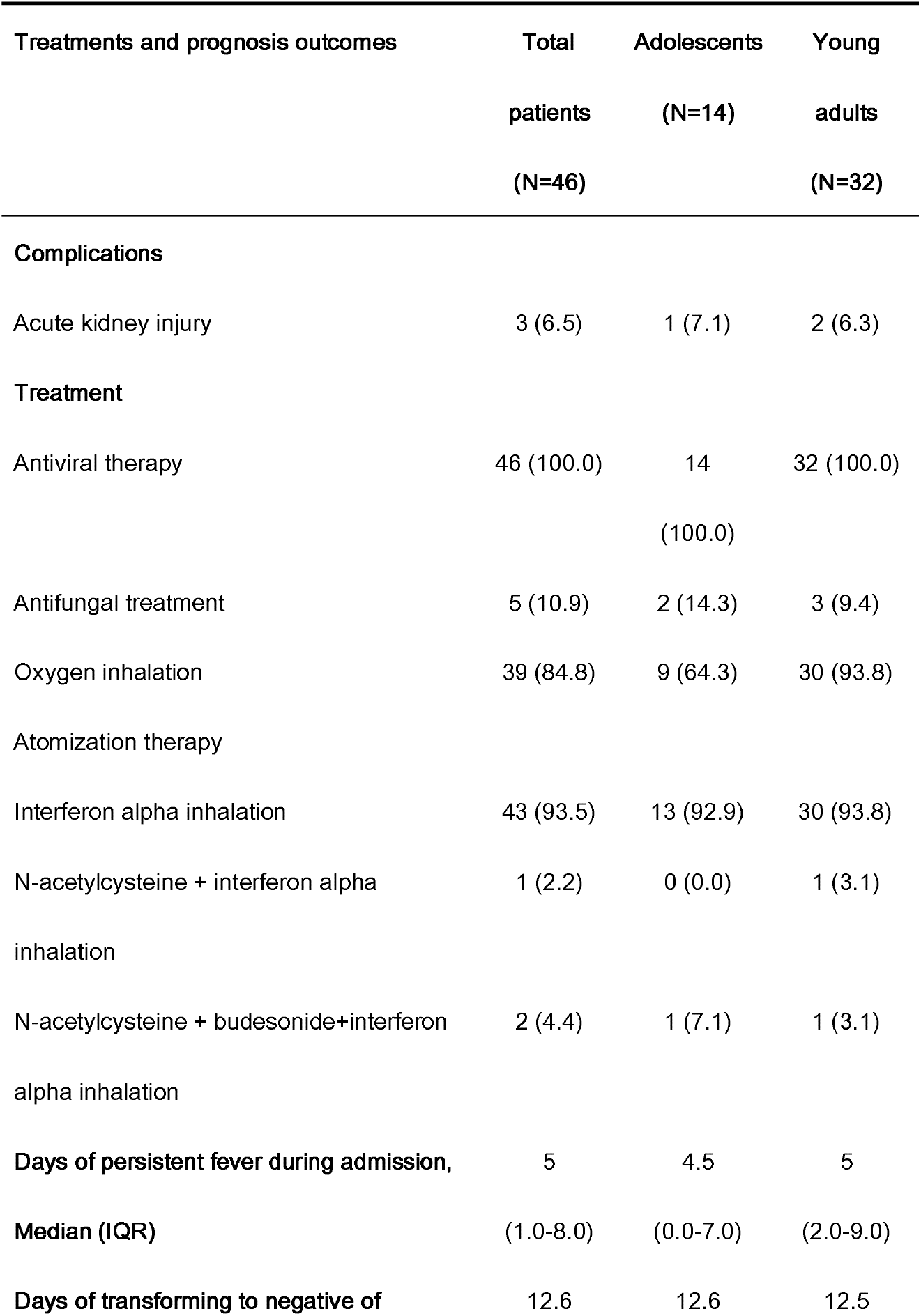
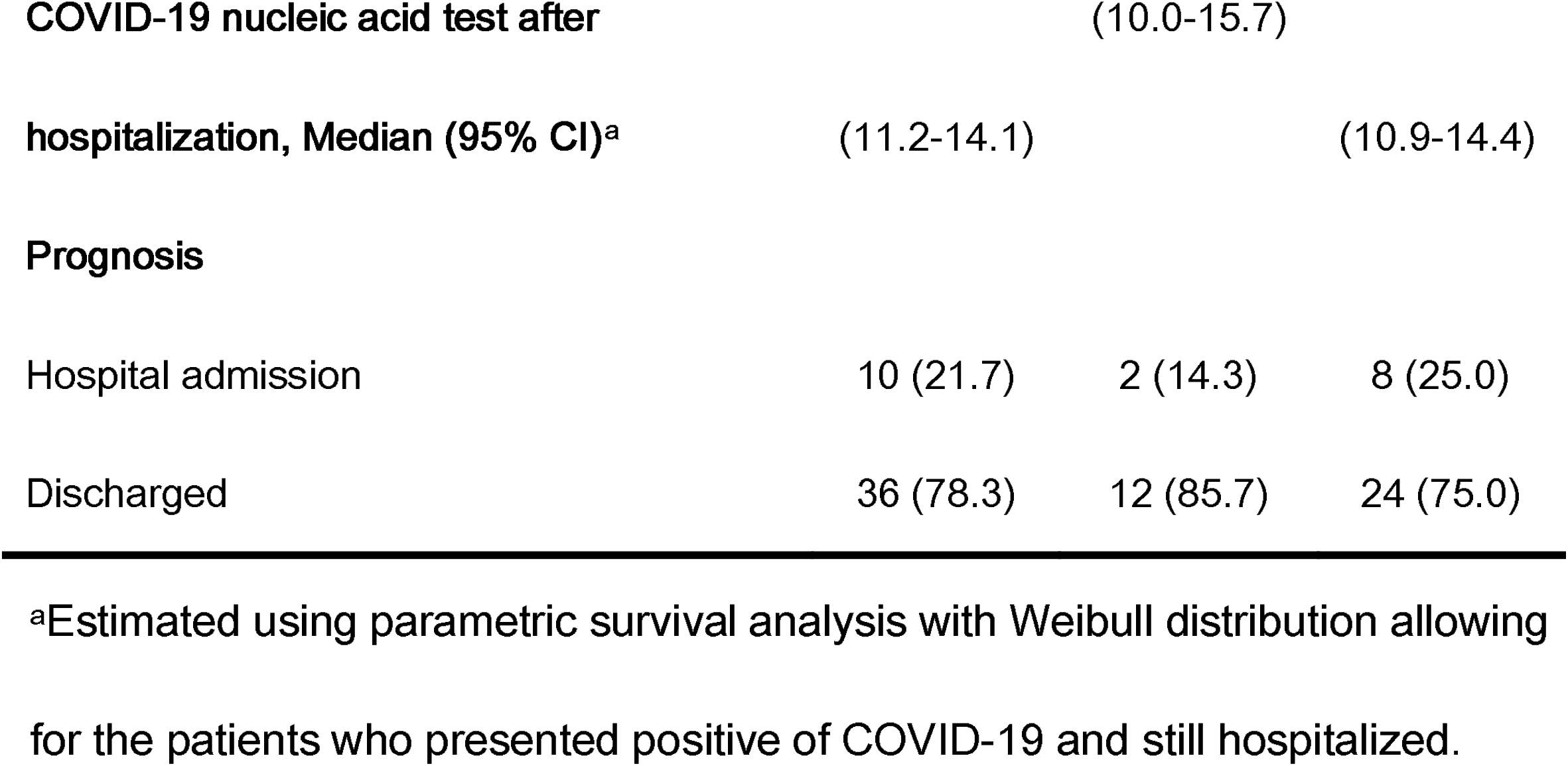
Treatments and prognosis outcomes in patients with COVID-19. Values are No. (%) unless stated otherwise.

We observed 4 asymptomatic cases at admission and both of them were consistently confirmed as asymptomatic cases by our face-to-face or telephone interviews. Their progresses of COVID-19 during treatment periods were shown in Figure 4. Two asymptomatic cases (case 2 and case 3) still did not show any symptoms until February 23, 2020. Asymptomatic case 1 have developed symptoms of shortness of breath, difficulty breathing, and chest tightness in 17 days after admission.

**Figure 4.**
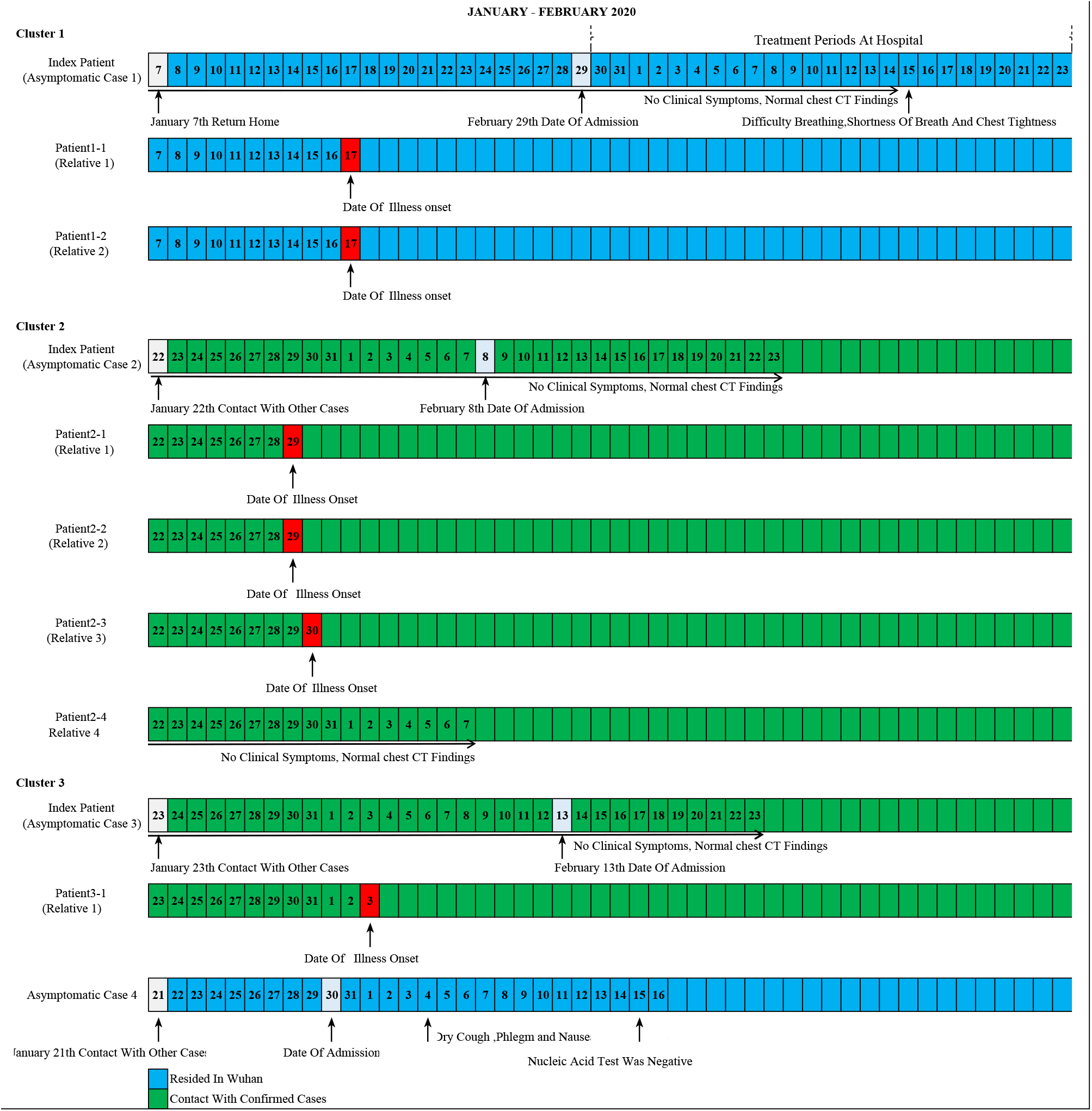
The progresses of clinical symptoms and lung CT findings during treatment periods for 4 asymptomatic cases and their family contactors. Numbers in boxes are calendar dates. The symptoms and the chest CT-findings related to COVID-19 were marked in onset dates by black arrows.

Asymptomatic case 4 have developed symptoms of dry cough, phlegm, and nausea in 6 days after admission and his COVID-19 nucleic acid have transformed into negative in 14 days after admission. We detected family-clustered events from 3 asymptomatic cases, which indicated that the transmission during their asymptomatic periods occurred between asymptomatic cases and their family close contactors. For example, two relatives of the asymptomatic case 1 who lived with him, and did not report other potential transmission sources, have developed illness in January 17, 2020.

## Discussion

This study, to the best of our knowledge, is the first to assess the epidemiological and clinical characteristics of COVID-19 in adolescent and young adult patients. We added new knowledge to deeply understand the characteristics of COVID-19 distributed in different sub-populations. We detected 4 asymptomatic cases out of 46 patients at admission. We reported a mean incubation period of 7.2 days in symptomatic cases, and could reach as long as 10 days with allowing for the truncated time periods of asymptomatic cases. We estimated a median serial interval of 1.9 days from the dates of illness onset in index patients to the date of developing illness in their family close contactors. We found that the most common symptoms were dry cough, fever, and expectoration. Only 29 (63.0%) of the patients showed the ground-glass opacity by chest CT scan. The typical changes of laboratory indicators were decreased white blood cell count, decreased lymphocyte count, decreased platelet count, increased lactate dehydrogenase, and elevated C-reactive protein. During the treatment, we found only 3 patients occurred acute kidney injury, and no other medical complications were reported. Nearly 80% of the patients were discharged in the end of follow-up.

The incubation period of COVID-19 in adolescent and young adult is longer than the elder patients. A retrospective study reported the mean incubation period was 5.2 days (95% CI: 4.1-7.0) and the 95^th^ percentile of incubation period was 12.5 days based on early COVID-19 patients from Wuhan ^2^. A later study, which used the data of travelers from Wuhan, estimated the mean incubation period to be 6.4 days (95% CI 5.6-7.7) and ranged from 2.2 to 11.1 days^15^. The similar studies reported a shorter incubation period (median = 4 days) for patients outside Wuhan^6 16^. However, most of these studies were based on the patients aged over 50 years. Knowledge gaps still persisted for the incubation period in younger COVID-19 patients.. In this study, we used patients with exact information for exposure time intervals and reported a mean incubation period of 7.2 days (95% CI 5.2-10.1) for patients aged under 35 years. The 95^th^ percentile of incubation period was 14.8 days (95% CI 10.4-22.00). With allowing for the right truncated periods for asymptomatic cases, the estimated incubation period (95^th^ percentile) could reach as long as 24.8 days (95% CI 14.9 - 47.6). Our findings highlighted the importance of extending the medical observing or quarantining time periods for adolescents and young adult of COVID-19 patients.

Our study suggests that the person-to-person transmission have occurred rapidly from adolescent and young adult infected cases of COVID-19 to their family contactors. We recorded 6 family-cluster events of COVID-19 in asymptomatic patients. We estimated the mean serial interval to be 6.5 days (95% CI 2.5 −17.4), which is shorter than that (7.5 days, 95% CI 5.3-19.0) estimated from early Wuhan patients ^2^. Most importantly, we estimated the median serial interval to be 1.9 days (95% CI 0.4 - 6.2), which was still lower than that (4.0 days, 95% CI 3.1-4.9) estimated in a recent modeling study^17^.

We provided evidence linking cluster-transmission to adolescent and young adult asymptomatic patients of COVID-19. In this study, four out of 46 patients were identified as asymptomatic cases. Three of them were identified as the primary cases for their contacting family members. Two asymptomatic primary cases were still show neither any symptoms nor chest CT findings during treatment. One asymptomatic primary case has suffered with difficulty and shortness of breath, and chest tightness during 17 days after treatment. However, all of their family close contactors have developed symptoms before the admission date of asymptomatic cases. Our findings were consistent with the existing evidence. Camilla Rothe et al. firstly reported an asymptomatic Chinese woman might be the transmission source for her two Germany business partners^18^. Zhen-Dong Tong et al. reported a 2-family cluster of COVID-19 patients in Zhejiang Province after each family’s primary case contacted with an asymptomatic case of COVID-19 from Wuhan^19^. Recently, a similar study has identified a 20-years age Chinese woman as an asymptomatic carrier who have infected her five family members^20^.

Compared with the early evidence from Wuhan patients, the adolescent and young adult patients of COVID-19 presented different patterns of symptoms and fewer abnormalities of laboratory indicators at admission. The most common symptoms were fever (83%), cough (82%), and shortness of breath (31%) in early elder patients from Wuhan^3^. Later studies with more case series reported other common symptoms including fatigue, gastrointestinal symptoms, upper airway congestion, myalgia, and headache^4 6 21^. The results of chest CT scanning indicated that nearly 80% of the early patients showed bilateral pneumonia, and ground glass opacity^3 4 21 22^. Laboratory examinations indicated that over 70% of the patients emerged lymphocytopenia, elevated lactate dehydrogenase, and elevated C reactive protein^3 4 23^. In this study, most common symptoms at admission were dry cough (81.0%), fever (69.1%), and expectoration (38.1%). Only 1 patients reported shortness of breath at admission. The proportion of reporting fever at admission decreased to 58.3% in adolescent patients. Nearly 60% of the patients showed ground-glass opacity changes by chest CT findings, which decreased to 50% in adolescent patients. Only 26.09% and 13.04% of all patients showed the bilateral patchy shadowing or consolidation by chest CT findings. By laboratory examinations, 63.0% of the patients had lymphocytopenia, which was close to the existing evidence. However, only a few patients had elevated levels of lactate dehydrogenase (19.6%), and C-reactive protein (19.6%). Both of these abnormalities of laboratory findings were less pronounced in adolescent patients.

Our study indicated that younger patients have better prognosis outcomes during the treatment.

Early studies reported that the nearly 40% of the patients have at least one medical chronic disease at admission and the common complications during the treatment included acute respiratory distress syndrome, shock, acute cardiac injury, arrhythmia, kidney injury, and liver dysfunction^3 4 6 24^. Most of the patients received antiviral therapy, and oxygen inhalation. Part of them received glucocorticoid therapy, or antifungal treatment. Nearly 20% of the patients were identified as severe cases and received mechanical ventilation and Extracorporeal Membrane Oxygenation (ECMO). In our study, only 1 (2.2%) patients were identified as severe cases at admission. After received antiviral therapy, interferon alpha inhalation, and oxygen inhalation, nearly 80% of the patients discharged at the end of the follow-up. Three patients developed severe kidney injury during treatment.

This study provided the initial evidence for the epidemiological and clinical characteristics of COVID-19 in adolescent and young adult. The longer incubation period indicated that longer time periods of medical observation and isolation are needed for suspected younger patients. The shorter serial interval indicated that the transmission could emerged rapidly between younger patients and their family members or close contactors. Compared with elder patients, younger patients had fewer typical signs and symptoms, and less abnormalities of laboratory findings. Fewer of them developed severe complications during treatment. Both of these evidence indicated that the adolescent and young adult might be the key subpopulation in the later stage for preventing the worldwide spread of COVID-19.

The study has some limitations. Firstly, we conducted this study only based on 46 patients, which enabled us to compare the epidemiological and clinical differences between adolescent and younger adult with significance tests. Our results needed to be replicated with large sample size. Secondly, at the end date of this study, nearly 20% of the patients still hospitalized, which limited us to fully illuminate the prognosis outcomes for the study patients.

## Conclusions

Compared with elderly patients, the adolescent and young adult COVID-19 patients had a longer incubation period, a shorter serial interval, and a higher odd to be asymptomatic. The transmission to their family close contactors occurred in several asymptomatic cases. Few of the study patients have developed complications during treatment.

## Data Availability

Data can be accessed by communcating with corresponding authors.

## Acknowledgment

We thank all the patients in the study and Huilin Liu, Feifei Yuan, Fang Yang, Xia Huang, Yingchun Jiang, Bangjuan Yu, Runze Deng, and Li Chen for their kindly help for data collection in the study hospital. This work was supported by the Fundamental Research Funds for the Central Universities (No.2020CDJYGRH-YJ03).

